# Is Recovery Just the Beginning? Persistent Symptoms and Health and Performance Deterioration in Post-COVID-19, non-hospitalised University Students - A Cross-Sectional Study

**DOI:** 10.1101/2023.10.20.23297203

**Authors:** Ashkan Latifi, Jaroslav Flegr

## Abstract

Many individuals experience persistent symptoms such as deteriorated physical and mental health, increased fatigue, and reduced cognitive performance months after recovering from COVID-19. Current data are limited on the long-term trajectory of these symptoms and their prevalence in milder cases. Our study aimed to assess the persistent effects of COVID-19 on physical and mental health, fatigue, and cognitive performance in a cohort of 214 students, averaging 21.8 years of age. Of these, 148 had contracted COVID-19 but were not hospitalized, with the time since infection ranging from 1 to 39 months. We utilized a comprehensive panel of cognitive tests to measure intelligence, memory, and psychomotor skills, and a detailed anamnestic questionnaire to evaluate physical and mental health. While contracting COVID-19 did not significantly impact overall health and performance, it was associated with increased reports of fatigue. However, the reported severity of the disease had a pronounced negative influence on physical health, mental well-being, fatigue, and reaction time. Trends of improvement in physical and mental health, as well as error rate, were observed within the first two years post-infection. However, fatigue and reaction time showed a trend of deterioration. Beyond the two-year mark, physical health and error rate continued to improve, while mental health began to deteriorate. Fatigue and reaction time continued to decline. Overall, our findings suggest that some effects of contracting COVID-19 can persist or even deteriorate over time, even in younger individuals who had mild cases that did not require hospitalization.

## 1. Introduction

December 2019 news shocked the world with reports of a novel viral disease emerging in China, known by the name COVID-19. A disease that was once reported to be mainly a respiratory infection has come to be known as an illness with a wide range of physiological, psychological, psychiatric, neurological, and anatomical negative impacts on the patients and survivors. A growing body of research is pointing to the post-acute sequelae of COVID-19 in the survivors. For example, depression and cognitive impairment (Lamontagne, Winters, Pizzagalli, & Olmstead, 2021), fatigue (González-Hermosillo et al., 2021), pain (Fernández-de-Las-Peñas, de-la-Llave-Rincón, et al., 2022), psychiatric and neurological complications, see (Zawilska & Kuczyńska, 2022) and (Santos, Rodríguez, Hernandez, & 2023), and lower health-related quality of life (HRQoL) compared with controls (Tsuzuki et al., 2022) are among the symptoms that these survivors may suffer from.

Studies also show that COVID-19 post-infection effects can negatively affect patients’ mental health. A systematic review and meta-analysis of COVID-19 survivors revealed that 50.1% of these survivors experienced at least one sequela for up to a year after being diagnosed with the infection. In addition, this meta-analysis reported that significant abnormalities on lung CT scan, abnormal pulmonary function, fatigue, psychiatric symptoms (primarily depression and PTSD), and neurological symptoms, e.g., cognitive deficits and memory impairment were observed in these survivors. In addition, this meta-analysis found that the elderly (specifically males) with a history of more severe forms of the disease and those with an underlying health or mental condition, moderated by the type and protocol of the treatment they received in the hospital, exhibited a higher risk of these sequelae. In terms of disease severity, subjects who experienced a severe form of COVID-19 had complications such as PTSD, cognitive deficits, concentration difficulties, sleep disturbance, and gustatory problems, whereas those with a history of mild COVID-19 developed high levels of anxiety and memory impairment (Zeng et al., 2023).

COVID-19 post-infection effects are also found to be associated with cognitive impairment in patients. In this regard, a systematic review pointed out that cognitive impairments were the most prominent in the domains of executive functions, attention, and episodic memory six months after disease onset in severe and moderate COVID patients (Tavares-Júnior et al., 2022). Another systematic review and meta-analysis also reported that subjects with a history of COVID infection experienced significant cognitive impairments compared with controls, however, only significantly in the sub-domains of processing speed and verbal memory but not attention, executive functions, fluency, visuospatial ability, and working memory (Houben-Wilke et al., 2022). The results of another recent meta-analysis and systematic review showed that COVID patients exhibited significant cognitive deficits compared with controls, corroborating the findings of former studies pointing to cognitive impairments in executive functions, working memory, attention, and processing speed in these patients (Sobrino-Relaño et al., 2023).

Regarding the studies reviewed so far, there are some points that need mentioning which justify the importance of future studies:

1) Few studies have considered the post-infection effects on those with mild to moderate COVID-19; in spite of the finding that even those with an asymptomatic COVID progression may still develop cognitive impairments (Amalakanti, Arepalli, & Jillella, 2021).
2) Few studies have investigated physical health, mental health, and cognitive functioning at once in the same sample, which is important because these domains are interconnected. For instance, cognitive impairment and depression are found to be pathophysiologically related (Papazacharias, Nardini, & 2012) and cognitive impairments are more abundant in individuals suffering from anxiety, depression, and bipolar disorder (Gualtieri & Morgan, 2008).
3) Few studies have systematically studied the effects of “time elapsed since the onset of infection” on physical health, mental health, and cognitive functions over a span of at least 2 years. Most of the studies done have considered the COVID-19 post-infection effects unfolding in less than a year from infection onset (Poletti et al., 2022); (Fernández-de-Las-Peñas, Martín-Guerrero, et al., 2022; Malesevic et al., 2023; Méndez et al., 2022).
4) The findings related to cognitive functioning, physical health complications, and mental health disorders associated with COVID-19 post-infection sequelae are varied and sometimes inconsistent. For example, although a study reported a prevalence of 82.3% for clinically significant levels of fatigue in the patients, another study found a prevalence of 11.5%; as to cognitive impairment/cognitive dysfunction, where a study reported a rate of 61.4%, another study put it at 23.5%; and as it regards depressive-anxiety symptoms the observed rate in one study was 23.5% and in another 9.5% (Sampogna et al., 2022).

Accordingly, the aim of the present study was to address the gaps identified in the existing literature concerning the post-infection consequences of COVID-19. The focus of our study was to explore the long-term effects of COVID-19. Specifically, we examined outcomes in cognitive functioning, physical health, and mental well-being over a period ranging from 1 to 39 months post-infection in individuals with mild to moderate cases of the disease. We concentrated on a cohort of 272 young, healthy university students. Data were collected through a medical history questionnaire, complemented by a broad panel of cognitive tests assessing aspects such as intelligence, memory, and psychomotor skills.

## 2. Materials and Methods

### 2.1. Study Participants

All students who took the online examination for an advanced course in evolutionary biology in 2022 and 2023 were invited to participate in an anonymous study “aimed at testing certain evolutionary psychology hypotheses and exploring the impact of various factors on exam performance.” Participants were informed about the voluntary nature of their participation and the scientific use of their data when they began the electronic questionnaire. They were also reassured of their ability to withdraw from the study at any time by simply closing the survey page.

Both the examination and the subsequent survey were facilitated separately on the Qualtrics platform. Upon concluding the exam, students were notified of their performance, i.e., the number of correctly answered questions. They were subsequently queried about this figure during the anonymous survey.

While the primary focus of the research was to evaluate the influence of diverse biological and social aspects on life history strategies, the project’s exploratory segment also committed to scrutinizing the impact of various factors on students’ performance in the exam and a set of cognitive tests. This was explicitly mentioned in the pre-registration form (DOI 10.17605/OSF.IO/FGRWD).

The online survey comprised several questionnaires and performance tests, of which only a few pertained directly to the study at hand. The project, inclusive of the informed consent acquisition method (achieved by clicking the designated button on-screen), received approval from the Institutional Review Board of the Faculty of Science at Charles University (No. 2021/19). The study adhered strictly to the relevant ethical guidelines for human subject research.

### 2.2 Questionnaires and tests

In this survey, we gauged the participants’ intelligence using the Cattel 16PF test (Variant A, Scale B) (Cattell, 1970) and their memory with a modified Meili test (Flegr et al., 2012; Meili, 1961). Initially, the Meili Memory Test involved presenting participants with a list of twelve distinct words (knife, handcuffs, pump, chain, tree, collar, ice, glasses, arrow, tank, bars, and rifle) for a period of 24 seconds. Approximately five minutes later, we prompted them for a free recall memory test, asking them to recollect as many of these words as possible. Subsequently, we presented them with a list of 24 words and asked them to identify the original twelve words in a recognition memory test.

We also evaluated the psychomotor skills of the participants, specifically their reaction time and precision, using two tests - the Choice Reaction Time Test and the Stroop Test. In the Choice Reaction Time Test, participants were directed to swiftly click with a computer mouse on a particular letter (A, B, C, or D) displayed on the screen. These letters were each assigned to one of four horizontally arranged buttons at the center of the screen. The button sequence was randomized for each of the six trials. We recorded the number of accurately selected buttons throughout the six trials (Choice Test Accuracy), along with the mean reaction time for these six trials (Choice Test Reaction Time).

Our variant of the Stroop Test included three distinct sections. Each section began only after students had received instructions and had time to rest, following the directive to start ‘when they are prepared’. In Part A, participants were required to select a specific word (e.g., “red”) from a set of four options (“red,” “green,” “blue,” “brown”), which were displayed in the center of the screen in a randomized order. These words were presented in a color that did not correspond to their actual meaning. The command specifying which word to choose was positioned at the top of the screen, with participants instructed to disregard the font color. Part B mirrored the conditions of Part A, but in this section, participants were required to select a word displayed in a specific color, whilst ignoring the words’ meanings. Part C was a slight variation of Part A, where the command specifying the word to be selected was consistently written in a contrasting color, not aligning with the meaning or color of the presented stimuli. Prior to each section, participants were provided with clear instructions, notified about the number of iterations (always five), and instructed to respond as rapidly as possible. Participants could initiate each section by clicking the ‘Start Test’ button. We recorded the number of correct responses across all 15 attempts (Stroop Test) and computed the average reaction time for all these attempts (Stroop Test Time). Additionally, we calculated the average reaction times for each of the three sections of the test (Stroop Test Time 1-3).

Participants were also tasked with solving three problems from the standard Cognitive Reflection Test (CRT) (Frederick, 2005). These problems were slightly modified to deter participants from looking up solutions online. The problems were: “Duckweed grows on the surface of a pond, doubling its area every day. If it takes 48 days for the duckweed to cover the entire surface of the pond, how many days did it take to cover half the surface?”, “Five workers can produce 5 parts in 5 minutes. How many minutes will it take for 100 workers to produce 100 parts?”, and “A car with a doll costs 110 CZK. The car is 100 CZK more expensive than the doll. How much does the doll cost?”. After solving each problem, participants were asked if they were already familiar with it. Approximately 18% of the participants recognized the problems, and their results were not included in the final assessment.

The ‘Reading Time’ variable was calculated as the mean Z-score of the time taken to read the instructions for all included tests. These instructions, concise in nature, were presented as short paragraphs on the webpage before each test. The ‘Error Rate Score’ was derived from the mean of the inverted Z-scores obtained from the Evolutionary Biology exam, IQ test, CRT, Choice Reaction Time test, and the Stroop test. The ‘Reaction Time Score’ was determined as the mean Z-score of Reading Time and reaction times captured during the Choice Reaction Time test, Stroop test.

In the anamnestic section of the questionnaire, participants were required to answer 19 questions related to their physical health. These questions covered the frequency of various conditions, including allergies, skin disorders, digestive tract disorders, metabolic disorders, common infectious diseases, orthopedic disorders, neurological disorders, headaches, physical pains, and other chronic physical issues. “Participants were also queried about their antibiotic usage over the past year and the preceding three years, their frequency of visits to a general practitioner, and any hospital stays that exceeded a week in the past five years. They provided responses using 6-point ordinal scales anchored by, e.g. ‘never’ and ‘daily or more frequently’. For the precise wording of all questions, refer to the questionnaire text attached to the preregistration form (DOI 10.17605/OSF.IO/FGRWD).

Further, they reported the number of non-mental health medications prescribed by a doctor that they were currently taking, with options ranging from 0 to 7, where 7 indicated six or more medications. Questions about their current physical feeling, usual physical feeling, and a comparison of their physical condition to that of their peers were answered using 6-point scales. Finally, participants were asked to estimate their life expectancy, with response options ranging from ‘more than 99 years’ to ‘less than 60 years.

The responses to these questions were inverted when a higher value indicated better health and a lower value indicated poorer health. The index of physical sickness score was then derived from the mean Z-scores of all these 19 questions (Flegr, 2021). Similarly, a mental sickness score was derived from participants’ responses to nine variables: frequencies of depression, anxiety, phobias, obsessions, other mental health problems, the number of prescribed mental health medications, and questions about their current mental state, usual mental feeling, and comparison of their mental condition to that of their peers, all of which were gauged using 6-point scales.

The fatigue index was calculated based on the mean Z-scores for five variables: the frequency of tiredness (6-point scale, anchored by ‘never’ and ‘daily or several times a day’), current level of tiredness, fatigue after returning from work/school, fatigue after several hours of bus travel, and fatigue after several hours of train travel (all using 6-point scales, anchored by ‘definitely not’ and ‘definitely yes’).

In the anamnestic section of the survey, additional demographic and medical history data were gathered from participants. These included their age and official sex as stated on their birth certificate (with men coded as 1 and women coded as 0), and history of SARS-CoV-2 (COVID-19) infection. Participants’ responses to their COVID-19 infection status were coded as follows: responses “not yet” and “no but I was in quarantine” were coded as 0 (COVID-19 -negative), “yes, I was diagnosed with COVID-19 “ was coded as 1 (COVID-19 -positive), while “probably yes, but I was not diagnosed with COVID-19 “ and “I am waiting for the result of a diagnostic test” were coded as NA (data not available), as these responses did not provide a definite confirmation of their COVID-19 status. Participants who confirmed their COVID-19 diagnosis were further asked to specify the number of months since the onset of their illness and to rate its severity on a 6-point scale (1: No symptoms, 2: Like mild flu, 3: Like normal flu, 4: Like severe flu, 5: I was hospitalized, 6: I was in ICU).

### 2.3. Data Analysis

To address potential issues arising from an unbalanced dataset (with women outnumbering men two-to-one and infected subjects similarly outnumbering their non-infected counterparts), irregularities in data distribution, and potential confounding variables, we utilized a non-parametric multivariate method for the analyses of the impact of COVID-19 infection status, severity, duration, and time elapsed since onset on health and cognitive performance. Specifically, we employed a partial Kendall correlation test, controlled for age, sex, and the survey year, to investigate the effects of three COVID-19-related variables. These tests, as well as t-tests and Chi^2^ tests used in the descriptive statistics section of the study, were conducted using the Explorer v. 1.0 R script (Flegr & Flegr, 2021), which leverages the ppcor R package (Kim, 2015). In the mixed-gender sample analysis, we controlled for age, sex, and the survey year (either 2022 or 2023). In the sex-specific analyses, we only controlled for age and year. The Kendall correlation test allows for the control of confounding variables and is robust against outliers and variable distribution shape in general. To adjust for multiple testing, we employed the Benjamini-Hochberg procedure, setting the false discovery rate (FDR) at 0.10 (Benjamini & Hochberg, 1995). The dataset for this study is publicly accessible on Figshare 10.6084/m9.figshare.24032700.

#### Technical notes

The term “effect” is used throughout the article in a statistical context to denote an observed association—the difference between the actual population parameter and the null hypothesis value. Only in the Discussion section do we differentiate between cause and effect. As the main part of the study has an exploratory nature, we discuss not only statistically significant effects but also trends that did not achieve formal significance.

## 3. Results

### 3.1. Descriptive Statistics

In total, 311 students participated in the evolutionary biology written examination, with over 95% of them consenting to partake in the subsequent anonymous study. Despite their initial agreement, some students either did not complete the questionnaire, or hastily clicked through it, providing uniform answers to a majority of the questions. The finalized dataset encompassed information on COVID-19 experiences from 272 individuals, representing 87.5% of the students initially approached for the study. In total, 54 people reported no prior COVID-19 infection, 152 had experienced the infection, 53 people possibly had COVID-19 but had not received a formal diagnosis, and 13 had not had the virus but had undergone quarantine.

From the dataset, we excluded one notably older individual, as well as 53 individuals who reported possibly having COVID-19, but without a laboratory-confirmed diagnosis. The final data set included 214 individuals, 66 who did not have COVID-19 and 148 (69.2%) who did. Among the 144 female students, 103 (71.5%) had experienced COVID-19, while among the 70 male students, 45 (64.3%) had; however, these differences were not statistically significant (Chi^2^ = 0.843, df = 1, p = 0.358). The average age of all students, female students, and male students was 21.77 (sd = 1.61), 21.81 (sd = 1.71), and 21.70 (sd = 1.39), respectively (Fig. 1). The difference in age between men and women was nonsignificant (t_(165.23)_ = 0.483, p = 0.630), as was the difference in age between those who had (21.84) and had not (21.62) experienced COVID-19 (t_(161.78)_ = −1.0085, p = 0.315).

**Fig. 1.**
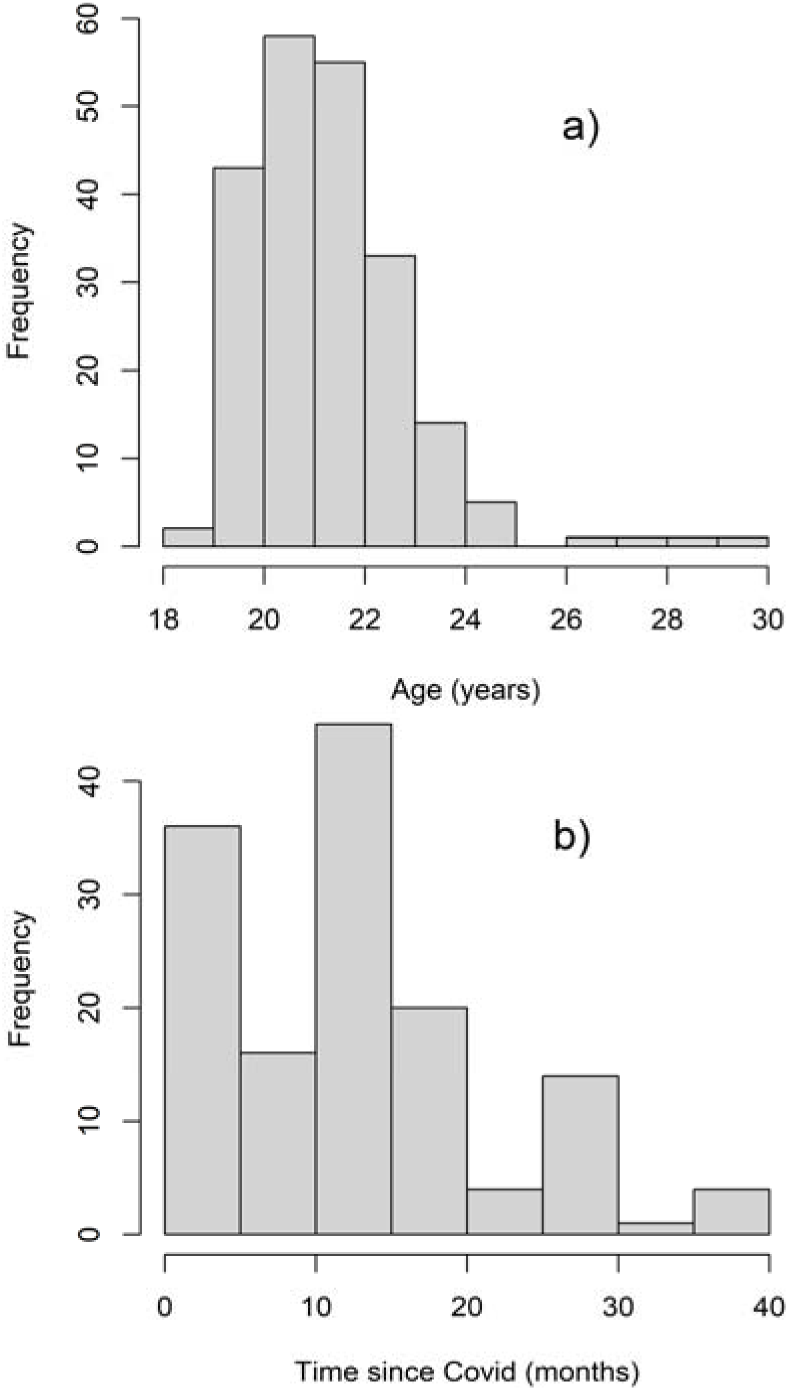
Distribution of age of participants and times since the beginning of COVID-19

Among the 148 subjects diagnosed with COVID-19, 15 (10.1%) reported “No symptoms”, 52 (35.1%) described it as “Like mild flu”, another 52 (35.1%) as “Like normal flu”, and 29 (19.6%) as “Like severe flu”; none reported being “hospitalized” or “in ICU”. Females reported a more severe course of COVID-19 than men (mean: 2.73 vs. 2.44, median: 3 vs. 2, Kruskal-Wallis Chi^2^_(1)_ = 4.04, p = 0.044).

The average time since the beginning of COVID-19 was 13.41 months (sd = 8.47), 13.31 (sd = 8.24) in women and 13.63 (sd = 9.05) in men; this difference was not significant (t_74.172_ = −0.197, p = 0.844) (Fig. 1). The descriptive statistics for the dependent variables related to health, and cognitive performance are presented in Supplementary Table S1.

The age of students did not exhibit any correlation with the likelihood of contracting COVID-19 (All: Tau = 0.024, p = 0.600, Women = 0.057, p = 0.315, Men: Tau = 0.058, p = 0.478) or the time elapsed since contracting the virus (All: Tau = 0.015, p = 0.788, Women: Tau = 0.010, p = 0.881, Men: Tau = 0.047, p = 0.659). On the other hand, it showed a positive correlation with the severity of COVID-19 (All: Tau = 0.185, p < 0.0001, Women: Tau = 0.195, p = 0.003, Men: Tau = 0.131, p = 0.212). Furthermore, a negative correlation was found between the reported severity of COVID-19 and the time elapsed since the infection (All: Tau = −0.121, p = 0.036, Women: Tau = −0.086, p = 0.215, Men: Tau = −0.123, p = 0.257).

### 3.2. Effects of COVID-19 exposure, the severity of COVID-19, and time since contracting COVID-19 on health and performance

The associations between COVID-19 exposure, the severity of COVID-19, and time since contracting COVID-19 with health, performance, and personality metrics are presented in Table 2 (partial Kendall Taus) and Supplementary Table S2 (p-values). Merely contracting the infection significantly influenced the fatigue status of all participants (i.e., the mixed sex group); however, this was not the case when participants were grouped by sex or when corrections for multiple tests were applied.

Figure 2 visualizes the impact of the severity of a COVID-19 infection on health, performance, and fatigue indices, broken down by gender. To provide context, the final two columns display these indices for individuals who have not contracted COVID-19. COVID severity consistently demonstrated significant effects, impacting the physical health of all participants (i.e., the mixed-sex group) and each sex group separately. Additionally, it significantly influenced mental health, fatigue status, and reaction times for all participants and females, but not for males.

**Fig. 2.**
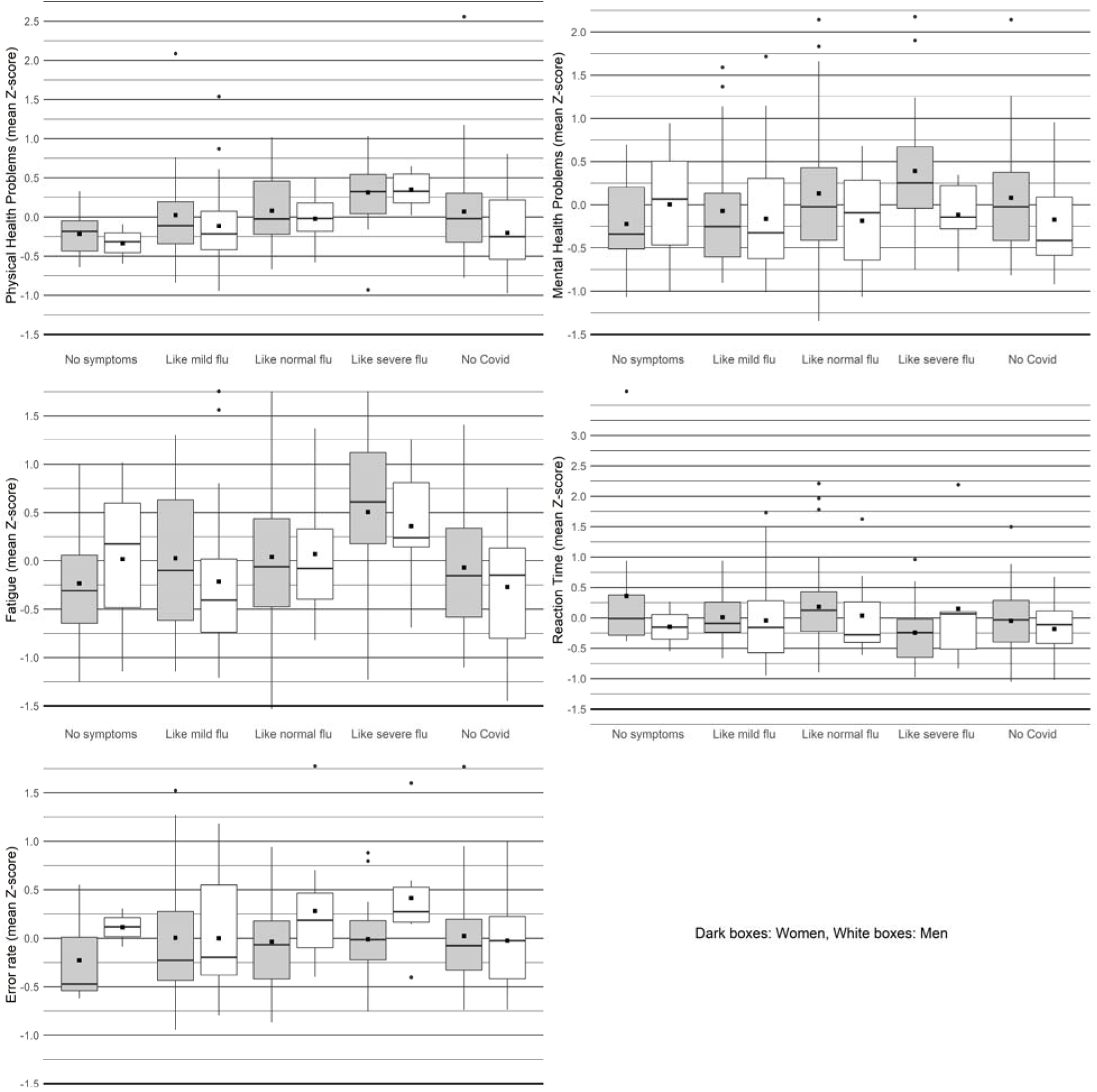
Effects of severity of COVID-19 (course) on five health and performance-related indices The figure displays boxplots representing the distribution of five health and performance-related indices across various categories of COVID-19 severity for both men and women. Each box encompasses the interquartile range (IQR), with a line inside the box indicating the median. The whiskers extend beyond the box to illustrate the range of variability (95% Confidence Intervals), and black squares denote the mean scores for each index.

In our study, the time elapsed since the onset of COVID-19 infection showed modest effects on both health and cognitive performance metrics. Specifically, we observed an improvement in physical health among males, while females experienced an exacerbation of fatigue levels. However, these changes were not statistically significant after applying the Benjamini-Hochberg correction for multiple testing. Consequently, during the three-year follow-up period post-infection, we did not detect the anticipated recovery, defined as the disappearance or substantial reduction of COVID-19 symptoms. Supplementary Figure S1 offers a potential explanation for these findings: the effects of COVID-19 appear to diminish in the first two years following infection, only to intensify subsequently. This pattern is particularly evident in the context of fatigue but is also noticeable in mental health outcomes and performance error rates. For an alternative interpretation of these observed trends, please refer to the discussion section.

To further investigate the pattern suggested in Figure 3—that the outcomes related to COVID-19 first show improvement and then deterioration after two years—we conducted separate correlation analyses for two different time frames: those who contracted the virus less than two years ago and those who contracted it at least two years ago. For individuals who contracted the virus less than two years ago, we observed non-significant decreases in the impact of COVID-19 on physical health (Tau = −0.04, p = 0.52) and mental health (Tau = −0.04, p = 0.54), and error rates (Tau = −0.05, p = 0.41). Simultaneously, a non-significant increase was observed in fatigue (Tau = 0.07, p = 0.24) and reaction time (Tau= 0.43, p = 0.49). In women, positive trends between the time elapsed since COVID-19 infection onset (less than 2 years) and physical heath (Tau =0.050, p = 0.51), mental health (0.044, p = 0.56), and reaction time (0.035, p = 0.63), with fatigue being significantly affected (Tau = 0.158, p = 0.039), and also a negative trend with error rates (Tau = −0.055, p = 0.46) were observed. In men, the indices of physical health (Tau = - 0.250, p = 0.036) and mental health (Tau = −0.250, p = 0.037) showed a significant decrease, fatigue and error rates demonstrated a negative trend (Tau = −0.083, p = 0.49, Tau = −0.056, p = 0.63, respectively), and reaction time proved a positive trend (Tau = 0.058, p = 0.62).

**Fig. 3.**
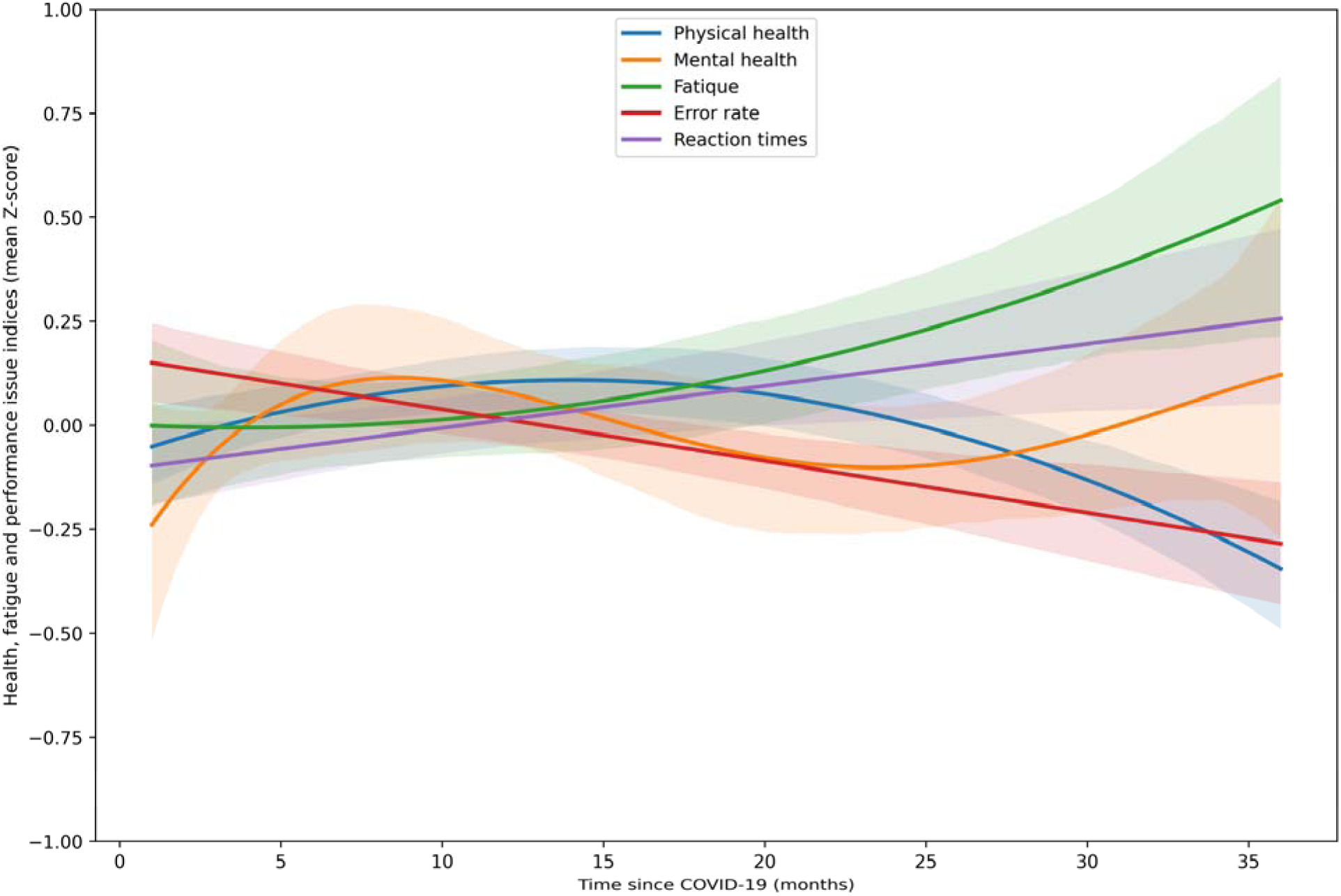
The correlation between the time since COVID-19 and five health and performance-related indices This scatterplot illustrates the relationships between five health- and performance-related variables and the time elapsed since contracting COVID-19. To visualize the trends, the points for each variable are fitted by polynomial curves, chosen based on comparisons of Adjusted R-Squared values. Specifically, a 2^nd^-degree polynomial curve is fitted for Physical Health, a 4^th^-degree polynomial curve for Mental Health, a 2^nd^-degree polynomial curve for Fatigue, a 1^st^-degree polynomial curve for Error Rate, and a 1^st^-degree polynomial curve for Reaction Times. (For the figure where data for all variables were fitted with the 3^rd^-degree polynomial curve, see Supplementary Fig. S1). Higher values on the y-axis indicate worse health and performance. The bands around the lines represent 80% Confidence Intervals (CI).

For the 22 subjects who contracted COVID-19 at least two years prior, all indices – with the exception of physical health, showing a negative trend (Tau = −0.223, p = 0.16) – demonstrated positive trends over the duration since their initial contraction of the virus – mental health (Tau = 0.094, p = 0.56), fatigue (Tau = 0.240, p = 0.15), error rate (Tau = 0.099, p = 0.55), and reaction time (Tau = 0.069, p = 0.67). In this regard, female participants demonstrated negative trends in physical and mental health (Tau = −0.244, p = 0.22, Tau = −0.030, p = 0.88, respectively).

Furthermore, trends were positive for fatigue (Tau = 0.140, p = 0.48), error rates (Tau = 0.010, p = 0.95), and reaction time (0.214, p = 0.28) in this group. Taking into account the male participants, the trends were pronounced for the improvement of physical health (Tau = −0.382, p = 0.28) and reaction time (Tau = −0.22, p = 0.53), as well as the deterioration of mental health (Tau = 0.27, p = 0.43), fatigue (Tau = 0.475, p = 0.24), and error rate (Tau = 0.052, p = 0.88). Given the small sample size of only seven men infected for more than two years, the absence of statistical significance was not surprising.

**Table 1.**
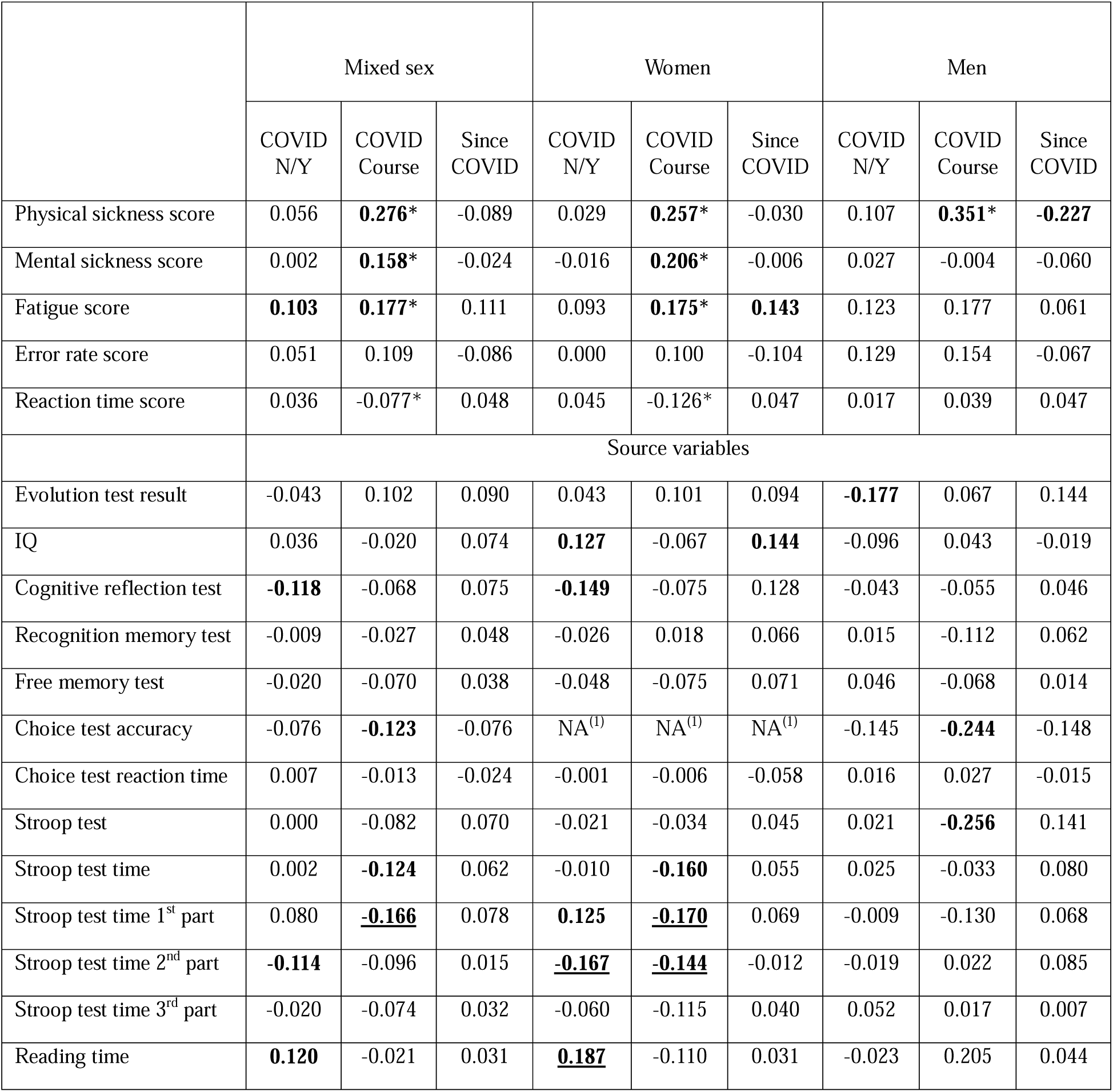
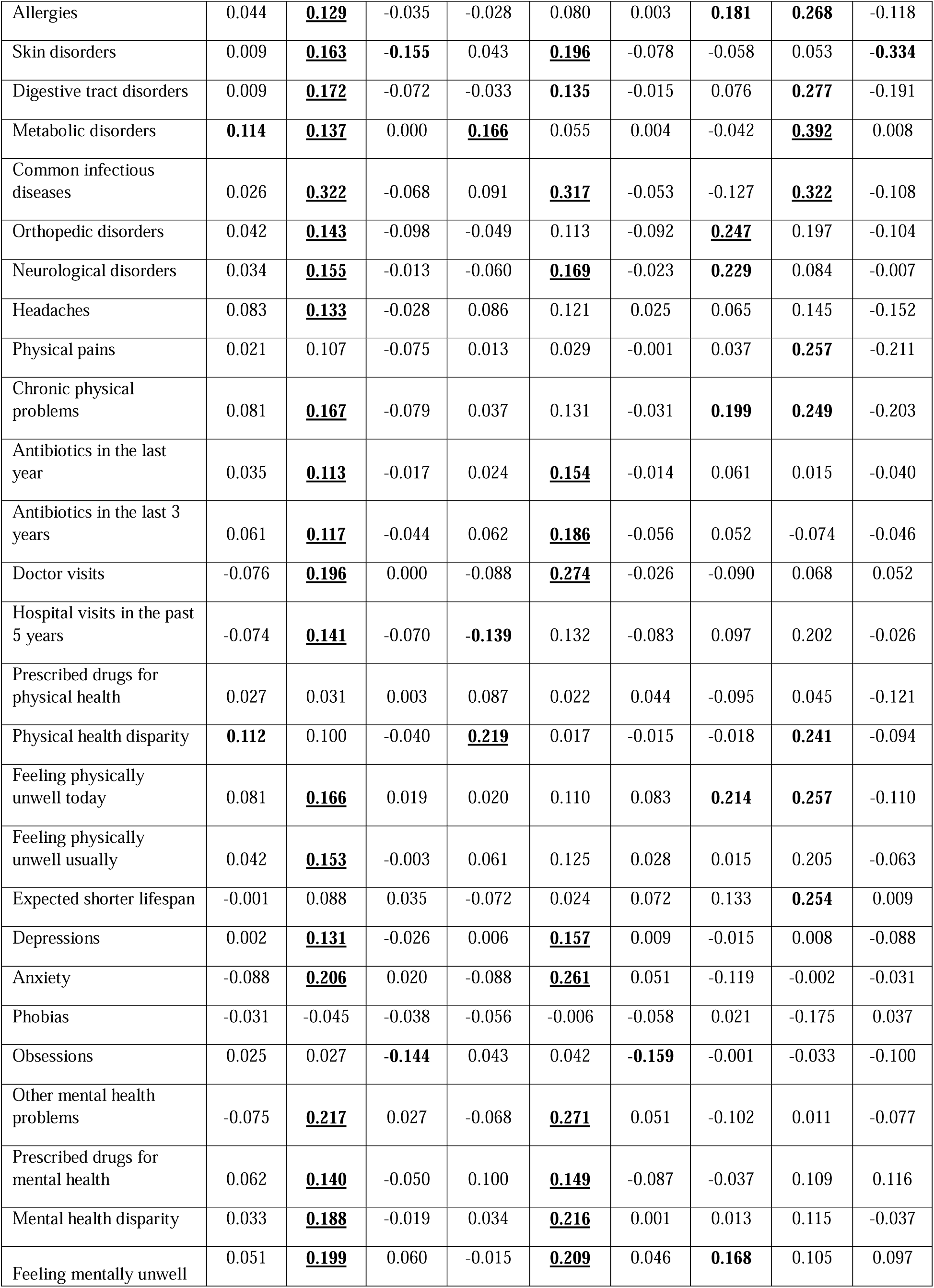
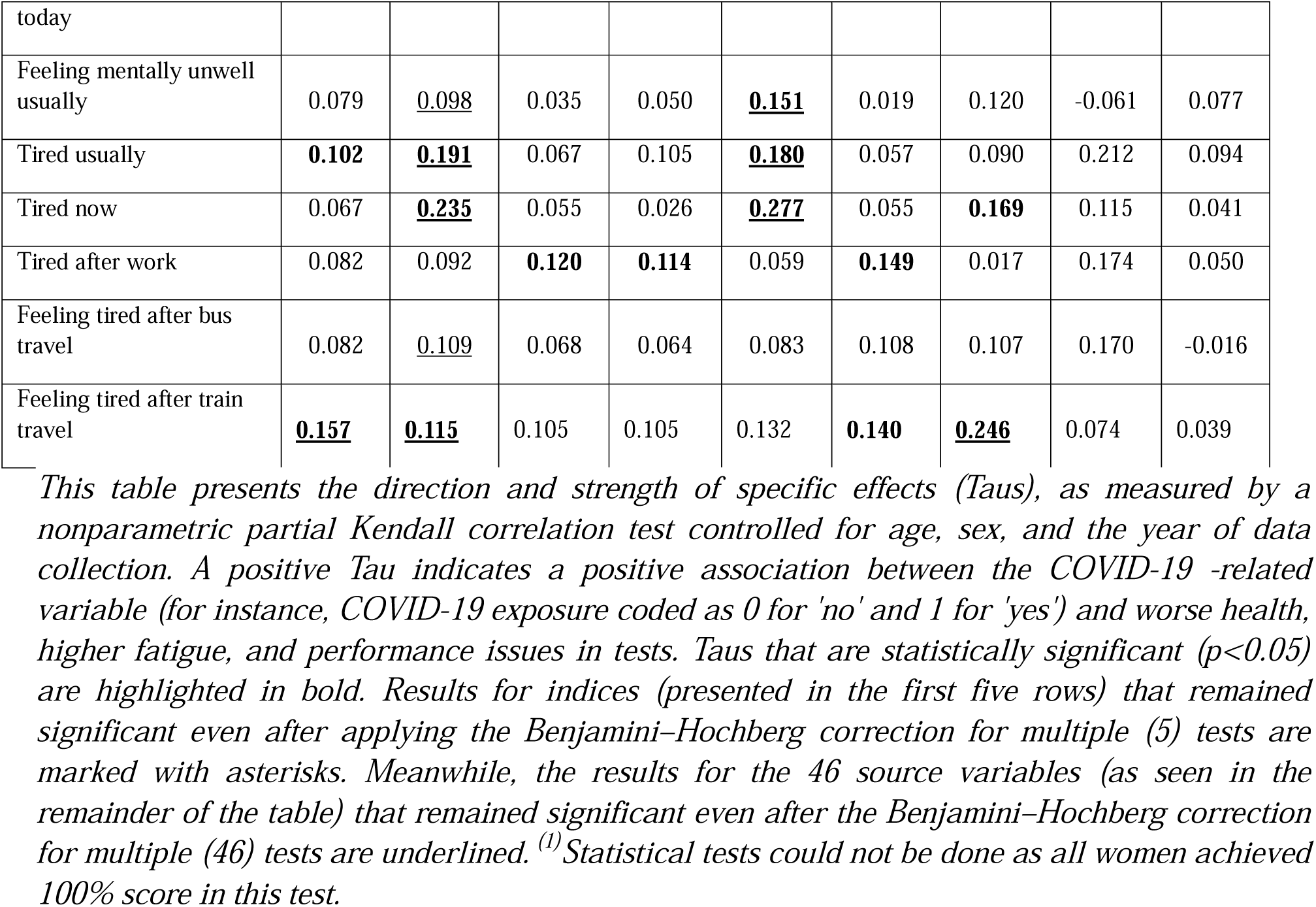
Effects of COVID-19 exposure, COVID-19 course, and time since COVID-19 on health, performance, and fatigue.

## 4. Discussion

Our study explores the enduring impacts of COVID-19 on health, fatigue, and cognitive performance among university students. In this population, having had COVID-19 did not significantly impact health and performance; however, individuals who contracted COVID-19 reported increased fatigue. On the other hand, the severity of COVID-19 significantly and negatively influenced physical health (in all, women, and men), mental health (in all and women), fatigue (in all and women), and reaction time (in all and women). The progression of all five indices under study relative to the time elapsed since COVID-19 infection suggests that trends of improvement in physical health, mental health, and error rate are observed for all participants in the first two years post-infection; however, fatigue and reaction time demonstrate trends of deterioration. Thereafter, physical health and error rate continue their marginal trend of improvement, while mental health worsens. Fatigue intensifies and reaction times extend (deteriorate) marginally. In the case of women, during the first two years post-infection, their physical health, mental health, and reaction time show a trend of deterioration, but error rate exhibits a marginal trend of improvement. It is noteworthy that fatigue significantly deteriorates for these participants in this period. Afterward, physical and mental health show a trend of improvement, fatigue and reaction time keep declining, and error rate marginally deteriorates. Nonetheless, for men during the first two years post-infection, mental and physical health significantly improve, fatigue and error rate demonstrate a trend of improvement, and reaction time deteriorates. Thereafter, physical health maintains a trend of improvement, but mental health, fatigue, and error rate show a trend of deterioration, and reaction time marginally improves.

The minimal or absent impact of merely contracting the infection on physical and mental health and cognitive performance, which contrasts with results of many already published studies, e.g. (Altuna, Sánchez-Saudinós, Lleó, & 2021; Delgado-Alonso et al., 2022; Flegr & Latifi, 2023; Havervall et al., 2021; Lamontagne et al., 2021; Lu et al., 2020; Marchi et al., 2023; Mazza et al., 2020; O’Mahoney et al., 2023; Zhao et al., 2020) could be related to the fact that all students included in our study were younger than 31 years old (95% were 20-24 years old). As is often the case with such a young population, most students experienced a relatively mild form of COVID-19; only about 20% described it as “severe flu,” and none of the study participants was hospitalized due to COVID-19. Furthermore, an observational bias could be present in our study: individuals who suffered severe cases might have discontinued or concluded their university studies and, as such, could not participate in our research. However, it is noteworthy that even in our young sample, enduring a case of COVID-19 that did not necessitate hospitalization still led to elevated fatigue levels, and trends of health and performance deterioration, although these trends were marginal and not statistically significant.

Our results align with previous studies that have identified a correlation between COVID-19 severity and subsequent declines in physical health (Flegr & Latifi, 2023; Han, Zheng, Daines, & Sheikh, 2022; Iqbal et al., 2021; Mizrahi et al., 2023), mental health challenges (Putri, Arisa, Hananto, Hariyanto, & Kurniawan, 2021; Shanbehzadeh, Tavahomi, Zanjari, Ebrahimi-Takamjani, & Amiri-Arimi, 2021; Zeng et al., 2023), and cognitive deficits (Ariza et al., 2023; Guo et al., 2022; Miskowiak et al., 2021). Collectively, these findings suggest that individuals with a history of more severe COVID-19 symptoms typically face more pronounced post-infection complications.

In our partial correlation tests, we did not observe a consistent change in health and performance symptoms over time following infection. The exceptions were a decrease in symptoms of impaired physical health among men and an increase in fatigue among women. This finding appears to contrast with anecdotal evidence and numerous published studies, which suggest that symptoms generally lessen with time since infection for most individuals (Poole-Wright et al., 2023; Tassignon et al., 2023), although in some cases they may stay constant or intensify (Carbone et al., 2022; Lucette et al., 2022; Qin et al., 2023). Visual inspection of Figure 3 and further analyses indicate that these inconsistencies may be attributed to a non-monotonic trajectory of changes as time since infection progresses, as well as to insufficient follow-up time after the illness. Some symptoms likely only emerged in the weeks and months after infection (COVID-19 broke out in 16 individuals 3 or fewer months before the study began). Other symptoms could decrease and disappear in the following months, only to reappear or start worsening more than 24 months after infection. However, it is important to keep in mind that our study had a cross-sectional design. This means it may be subject to a cohort effect, where individuals infected longer ago were exposed to different virus variants than those infected more recently. Thus, the time since infection can overlap with the specific variant of the virus that caused the infection. It is also likely that, in Czechia, young individuals infected in 2020 and the first half of 2021 had not received the COVID-19 vaccine prior to contracting the virus. Therefore, stronger symptoms in individuals with a very long time interval since infection may not be the result of gradual symptom intensification over time but rather the result of individuals being infected with different, more virulent, SARC-CoV-2 variants more than two years ago and not being vaccinated before getting sick. However, it is important to note that the negative correlation we observed between the time elapsed since COVID-19 and the reported seriousness of the course of the disease argues against this explanation.

Post hoc analysis on the effects of COVID-19 on specific variables used for index computation unveiled intriguing differences across genders. Following infection, men tended to score lower on the evolutionary biology knowledge test, whereas women exhibited poorer performance on the Cognitive Reflection Test. A unique aspect of our research was the inclusion of the Reading Time test, which measured the speed at which students read the instructions for each test. This was the only test where students were not explicitly aware they were being tested. As such, factors like competitiveness were less likely to influence the results, while urgency or haste may have played a role. In this test, women who had contracted COVID-19 took noticeably longer to read the instructions compared to their non-infected counterparts. No such trend was observed in men. However, among men, an increase in reading time was associated with a more severe COVID-19 experience; this association approached statistical significance (Tau = 0.205, p = 0.053). No significant association was observed between memory test outcomes and the examined COVID-19-related variables for all participants, males, or females.

Our results were in agreement with the findings of the studies that reported female COVID-19 patients to be more likely to develop long COVID-19 attributed cognitive deficits (Godoy-González et al., 2023; Jiménez-Rodríguez et al., 2022; Lippi, Sanchis-Gomar, & Henry, 2023) and in disagreement with a study in which being male was associated with more cognitive impairment (Hartung et al., 2022); however, only 1% of Hartung et al.’s participants demonstrated moderate cognitive impairment in comparison with the 26% who exhibited mild cognitive impairment. The reason for these sex-related differences may in part be due to male and female patients’ different immunological response to the infection. As a study showed, male COVID-19 patients had higher plasma levels of innate immune cytokines; however, female COVID-19 patients had more robust T-cell activation in the acute phase of the disease (Takahashi et al., 2020). It is possible that the more robust activation of T-cells can result in longer and more durable neuroinflammation, which in turn could lead to higher cognitive impairment in females.

Interestingly, our data showed that women who experienced more severe COVID-19 symptoms demonstrated faster reaction times in the Stroop Test. This effect did not appear even remotely in a simpler Choice Reaction Time Test, where participants were only required to click a specific button on the screen out of four possible choices. Women who had recovered from an infection also scored higher in a concise 12-item intelligence test. While these effects were not observed in men, those who had contracted COVID-19 made fewer errors on the evolutionary biology test compared to men who hadn’t been infected.

These intriguing results could possibly be linked to a ‘resilience effect,’ where the process of overcoming a substantial health hurdle might unintentionally boost certain cognitive abilities (Flood & Keegan, 2022). Prior research has demonstrated, for instance, that mild stress can improve performance on non-declarative memory tests (Hidalgo et al., 2012). Additionally, stress may have an adverse effect on the personality trait of conscientiousness, inclusive of chronic infection-related stress (Lindová, Příplatová, & Flegr, 2012), which adversely impacts the personality trait of conscientiousness (Zhu et al., 2022). This is relevant as high conscientiousness has been found to negatively influence performance on specific cognitive tests (LePine, Colquitt, & Erez, 2000), possibly because of a tendency toward overcaution or overthinking. The observation in our study of the positive impact of having experienced COVID-19 on women’s performance in an intelligence test and men’s performance in an evolutionary biology examination necessitates further exploration, given that a variety of confounding factors could have potentially influenced this outcome.

Regarding health-related variables, women who had experienced COVID-19 reported a higher prevalence of metabolic diseases and perceived their physical health as inferior compared to their peers. Simultaneously, these women reported fewer hospital visits. This observation might be attributed to the artifact of conducting multiple tests, as setting the False Discovery Rate (FDR) to 0.1 means that 10% of the positive results are expected to be false positives. Men who experienced COVID-19 recounted a more frequent occurrence of allergies, orthopedic issues, neurological problems, and other long-term physical conditions, and they also reported feeling unwell, both physically and mentally. While all of these effects were comparatively strong, only the influence of COVID-19 on orthopedic issues retained its significance even after accounting for multiple testing. Accordingly, examining the source variables results corrected for type I errors, our findings regarding ‘orthopedic disorders’ in males and ‘metabolic disorders’ and ‘physical health disparity’ in females were in agreement with those studies that observed deteriorated physical health in these areas in post-COVID19 patients (for reviews on orthopedic disorders see (Slouma, Abbes, Kharrat, & Gharsallah, 2022) and (Slouma et al., 2023) and for a review on metabolic disorders see (Steenblock et al., 2021). With regard to cognitive functions, our study’s findings on females’ reading time scores are consistent with earlier studies that have found an association between COVID-19 infection and impairment in information processing speed (Almeria, Cejudo, Sanz-Santos, Deus, & Krupinski, 2023; Ferrucci et al., 2021; Flegr & Latifi, 2023). However, unlike these studies, we found a significant negative correlation between COVID-19 infection and females’ reaction times in the Stroop test, indicating improved reaction times compared to those who were not infected. The reason for this latter finding remains to be understood; nevertheless, there is a pattern in our findings that points to a hypothetical explanation. As our female participants progressed through the Stroop test, their reaction times showed notable improvement. In contrast, the male participants exhibited a slight but discernible trend toward slower reaction times. This suggests that performance in reaction time tests is influenced by learning (which predominates in women) and fatigue (which predominates in men). As a result, the magnitude and direction of COVID-19’s impact on reaction times hinge on the duration of the specific test employed in a study and the proportion of men and women in the sample examined.

Across the entire cohort, the severity of COVID-19 exhibited a significant correlation with almost all physical and mental health-related variables, even after adjusting for multiple tests. For women, a strong correlation was noted with the incidence of common infectious diseases and the frequency of antibiotic use (both being proxies for immune deficiencies), the number of visits to the general practitioner, frequency of skin diseases, neurological diseases, frequency of anxiety and depression, other mental health issues, the amount of medication currently being taken for mental disorders, how they rated their health in comparison to their peers, and how they currently felt mentally unwell.

In men, the severity of COVID-19 showed a particularly robust correlation with the frequency of metabolic disorders (Tau = 0.392) and with the frequency of common infectious diseases (Tau = 0.322). However, it also significantly correlated with allergies, gastrointestinal diseases, the frequency of physical pain experiences, and the frequency of other long-term physical issues. Men with more severe COVID-19 also rated their physical health as worse when compared to their peers, reported feeling physically unwell, both in the present and typically, and predicted their lifespan to be shorter. Although no correlation reached statistical significance for men’s mental health (as the sample included only 45 men who had contracted COVID-19), certain correlations were nevertheless relatively strong (with Taus > 0.1). Specifically, the severity of COVID-19 in men demonstrated a notable correlation with the number of various types of medications currently being taken for mental health issues, their comparison of mental health issues to those of their peers, and their present state of feeling mentally unwell.

Over time following infection, there was a general improvement in health status across nearly all variables. However, the correlations between health-related variables and the time elapsed since infection were relatively low and not statistically significant. For men, these negative correlations were stronger, particularly in relation to skin problems (Tau = −0.334), but also allergies, gastrointestinal complications, orthopedic issues, the frequency of common infectious diseases, headaches, other physical discomforts, and other long-term physical problems. The number of different types of medications currently taken for physical problems, how they rated their physical health compared to their peers, and how they felt physically unwell both today and usually, all declined with time since COVID-19. On the contrary, positive correlations emerged for some mental health-related variables, indicating a potential increase in issues over time from infection. This was the case for the number of types of prescribed medications currently taken for mental problems, feeling mentally unwell today, and usually feeling mentally unwell. Despite the relative strength of these trends, none reached statistical significance among men. Among women, these trends were weaker, and for three variables, namely phobias, the number of different types of medications currently taken for mental problems, and especially obsessions, the values even decreased over time since COVID-19. In this regard, our findings are aligned with studies that found a trend of improvement in the physical health of post-COVID-19 patients over time, e.g., (Oliveira, Jason, Unutmaz, Bateman, & Vernon, 2023). Deterioration of mental health conditions over time in post-COVID-19 patients is also reported in earlier studies (e.g., (Colizzi et al., 2023; Houben-Wilke et al., 2022)-.

All the source variables for fatigue positively correlated with having had COVID-19, the severity of COVID-19, and the elapsed time since infection. The strongest correlation was observed with the severity of COVID-19, wherein the relationship was significant for four out of the five examined variables. The most prominent correlation was the response to the question of how tired the participant feels at the moment (Tau = 0.235, women: Tau = 0.277, men: Tau = 0.115). For men, the strongest relationship was observed between having had COVID-19 and experiencing fatigue after a long train journey (Tau = 0.246) and feeling tired at the present moment (Tau = 0.169). Perhaps the most concerning finding was that fatigue does not diminish but intensifies over time since having had COVID-19. This upward trend was statistically significant in the case of fatigue experienced after work (for all, and women) and after a long train journey (for women). Our results diverged from those studies that reported decreasing levels of fatigue over time in COVID-19 patients (Fumagalli et al., 2022; Steinmetz et al., 2023; Van Herck et al., 2021). However, they align with the findings of a study that observed a trend of increasing fatigue over the months 1, 3, 6, and 12 following the onset of COVID-19 infection (Mazza et al., 2022). The discrepancies in findings could result from differences in study design and, importantly, variations in the duration over which the respective changes were monitored. For instance, one study noted an inverse trend in fatigue levels related to disease severity when comparing two assessments of the same COVID patients conducted approximately four months apart. While the initial assessment showed a positive correlation between COVID severity and fatigue, the follow-up indicated a negative correlation (Peterson, Sarangi, & Bangash, 2021). Another study documented a nonlinear progression of fatigue levels throughout the disease’s trajectory: fatigue peaked during its acute phase, then decreased and stabilized around months 5 and 9, only to rise again at month 12 (Seeßle et al., 2022).

Students on the higher end of our age spectrum reported a more severe course of the disease. This was rather unexpected, considering the relatively young age of all students (all under 31 years old) and the narrow age range of the study participants. This correlation was stronger (Tau = 0.195) and significant among female students. The correlation was not significant among male students. Nevertheless, even in the case of men, the Kendall Tau value was observed to be 0.131, which corresponds to a Pearson’s r value of 0.16. This is generally considered a moderate correlation in the context of biopsychological research, rather than a weak one.

### 4.1 Strengths and Limitations

A key strength of our study is the comprehensive and representative sample of biology students in Prague. The high participation rate ensures that our findings accurately represent this specific group. Moreover, the sample’s homogeneity, stemming from the shared academic and likely socio-economic backgrounds of the students, minimizes variability in potential confounding variables, enhancing the study’s analytical precision. This uniformity also allows even subtle effects to be more discernible. However, this same homogeneity does pose a limitation: it narrows the scope of our findings’ applicability. We will delve deeper into the implications of this limitation later in our discussion.

Importantly, participants were kept unaware of the study’s focus on COVID-19, not only at the outset but throughout the questionnaire’s duration (which received IRB approval). This strategy of incorporating COVID-19-related questions into the survey without explicit disclosure significantly reduced response bias tied to pre-existing attitudes or beliefs about the virus. Moreover, the wide range of topics covered in the study would have made it unlikely for participants to deduce that COVID-19 was a key focus. These measures ensure the accuracy and representativeness of our data, thereby enhancing the validity of our findings.

Finally, our study considered the time elapsed since disease contraction, an element frequently neglected in similar research. This enabled us to delve into the potential long-term effects of COVID-19, providing valuable insights for shaping post-recovery care strategies.

Indeed, this study also bears certain limitations. First, while the cognitive test performances were objectively measured, our reliance on self-reported data specifically for participants’ health status could introduce inaccuracies due to recall bias or subjective perceptions. Participants’ recollections of their symptoms and their personal assessments of health may not perfectly reflect their actual medical conditions, potentially skewing those aspects of our results.

Secondly, the cross-sectional design of this study complicates the differentiation between the effects of time since contracting COVID-19 and potential cohort effects. These could be influenced by the evolution of the SARS-CoV-2 virus and the succession of its variants in the human population. Furthermore, this design challenges our understanding of causality. At first glance, the observed correlation between the time since infection and fatigue might appear primarily as a result of either the virus’s cumulative impact or the cumulative effect of organ damage caused by the virus during the COVID-19 illness. However, it is also conceivable that both the time since infection and fatigue might be influenced by a third variable, such as the evolution of virus strains. Hence, future longitudinal studies are an absolute necessity to conclusively establish any causal relationships.

While the observed effects were statistically significant, particularly in the combined sample of both genders, the effect sizes might seem relatively small. Kendall Tau values of 0.1, 0.2, 0.3, and 0.4 correspond to Pearson’s r values of 0.16, 0.31, 0.45, and 0.59, respectively. Based on the traditional Cohen classification, most of the observed effects fall into the “weak” category, and only those with a Tau greater than 0.26 could be classified as “moderate.” However, it is critical to remember that the impacts of biological factors on performance and personality tend to be relatively modest. For instance, a similar study investigating the effects of COVID-19 and vaccination on performance and health in a larger but more heterogeneous online population found effects to be typically 3-5 times weaker, albeit still significant due to the large sample size (Flegr & Latifi, 2023). The generally small effect sizes of biological factors on, e.g., cognitive performance observed in cross-sectional studies within the human population can be attributed to the presence of numerous confounding variables beyond the primary independent variable (in this case, SARS-CoV-2 infection). These confounders, such as cooperativeness, competitiveness, and anxiety, influence the dependent variables (in this case, intelligence, reaction rate, fatigue etc.). Consequently, they reduce the proportion of variance in the dependent variable’s outcomes that can be attributed to the independent variable, thereby diminishing the observed effect size.

Furthermore, the sample for our study, predominantly consisting of university students from one of the top Czech universities, does not represent the broader population. This demographic limitation restricts the generalizability of our findings, thus underlining the importance of conducting similar research using diverse and independent samples to reinforce the validity and reliability of our observations.

## 5. Conclusions

The principal revelation of our study is the acknowledgment that the repercussions of contracting COVID-19 can persist for an extended period and may even intensify over time, even among younger individuals who are generally considered resilient to the virus. The probability and extent of these lingering effects correlate with the severity of the initial COVID-19 infection, including in individuals whose case was mild enough not to require hospitalization.

While the physical health sequelae of COVID-19 tend to diminish within the first three years following infection, this trend does not apply to all consequences of the virus. One of the most significant findings from our study is that fatigue levels progressively increase with time elapsed since infection during the first three years, i.e., across the entire period covered by our study. Consequently, it seems probable that fatigue is not merely a result of a general and transient health deterioration, but rather a specific yet previously uncharacterized manifestation of COVID-19.

In summary, our study underscores that many critical aspects of the pandemic, especially the long-term effects of the disease, remain inadequately researched and should warrant far greater scientific focus than currently accorded.

## Data Availability

The dataset for this study is publicly accessible on Figshare 10.6084/m9.figshare.24032700.

https://10.6084/m9.figshare.24032700

## Acknowledgment

This research was supported by Czech Science Foundation, grant number 22-20785S. Our sponsor had no involvement in the study design, the collection, analysis and interpretation of data, the writing of the report, or in the decision to submit the article for publication.

## Author contributions

JF planned the study and collected data, JF and AL analysed the data and wrote the article.

## Data availability

All data are available in the public repository figshare 10.6084/m9.figshare.24032700

---

**Supplementary Table S1.**
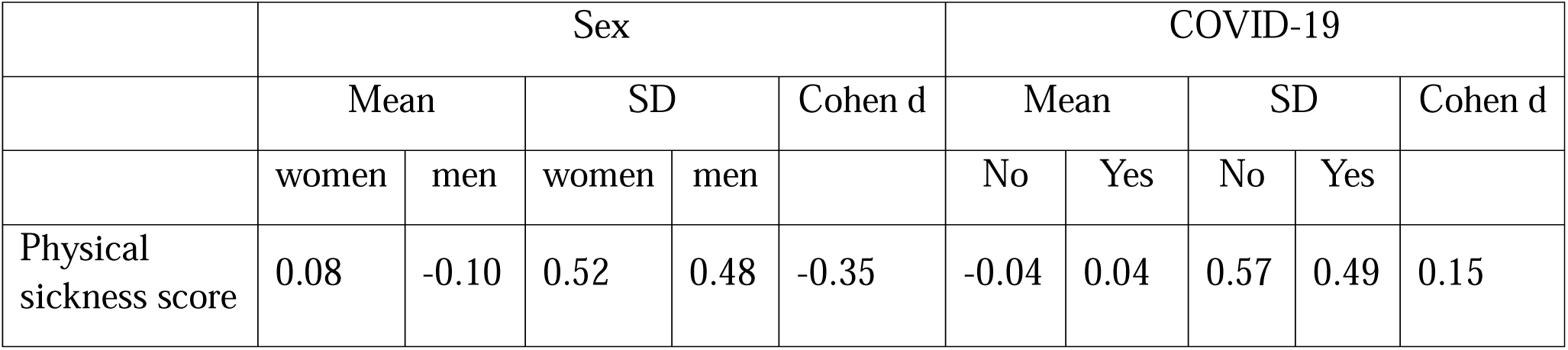
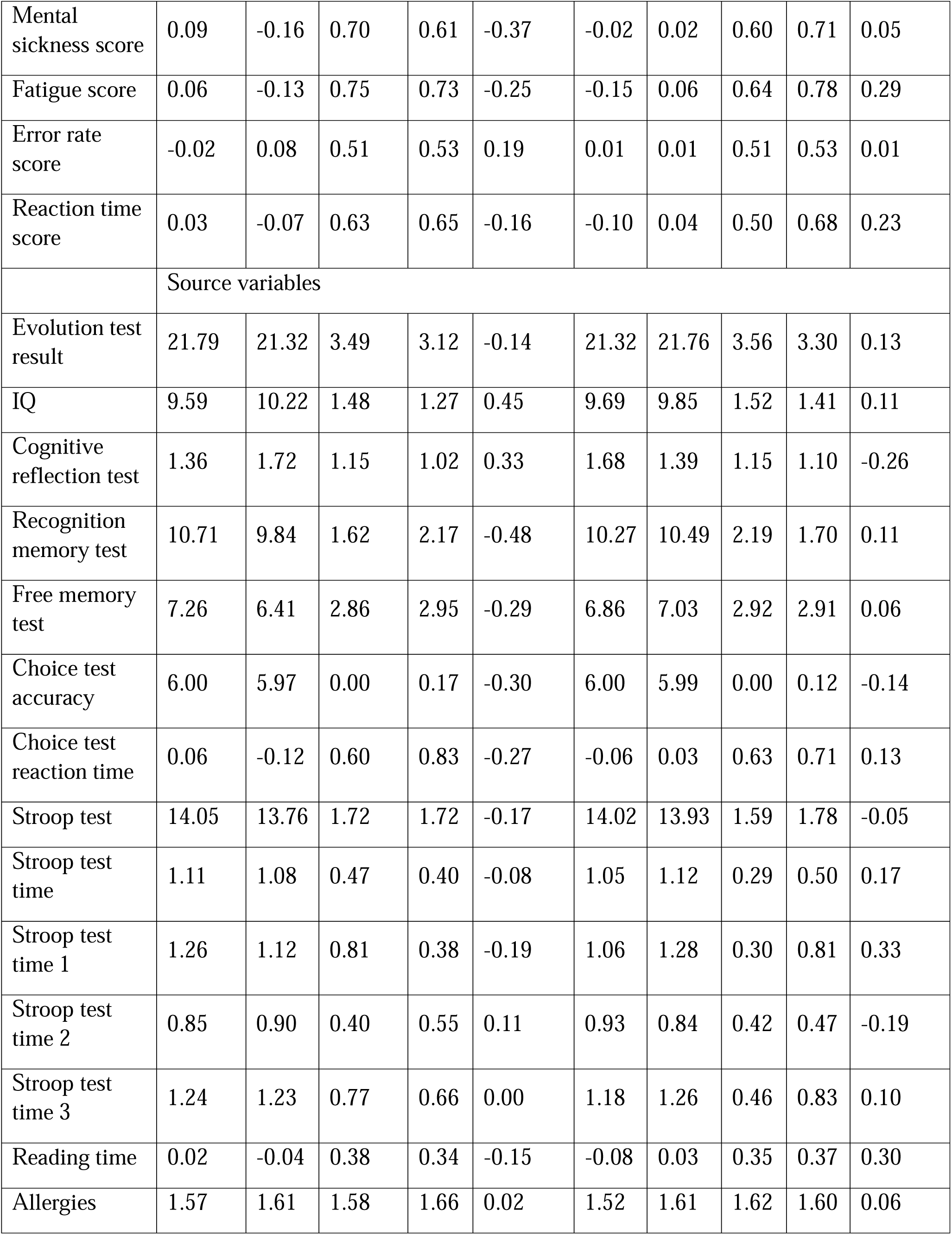
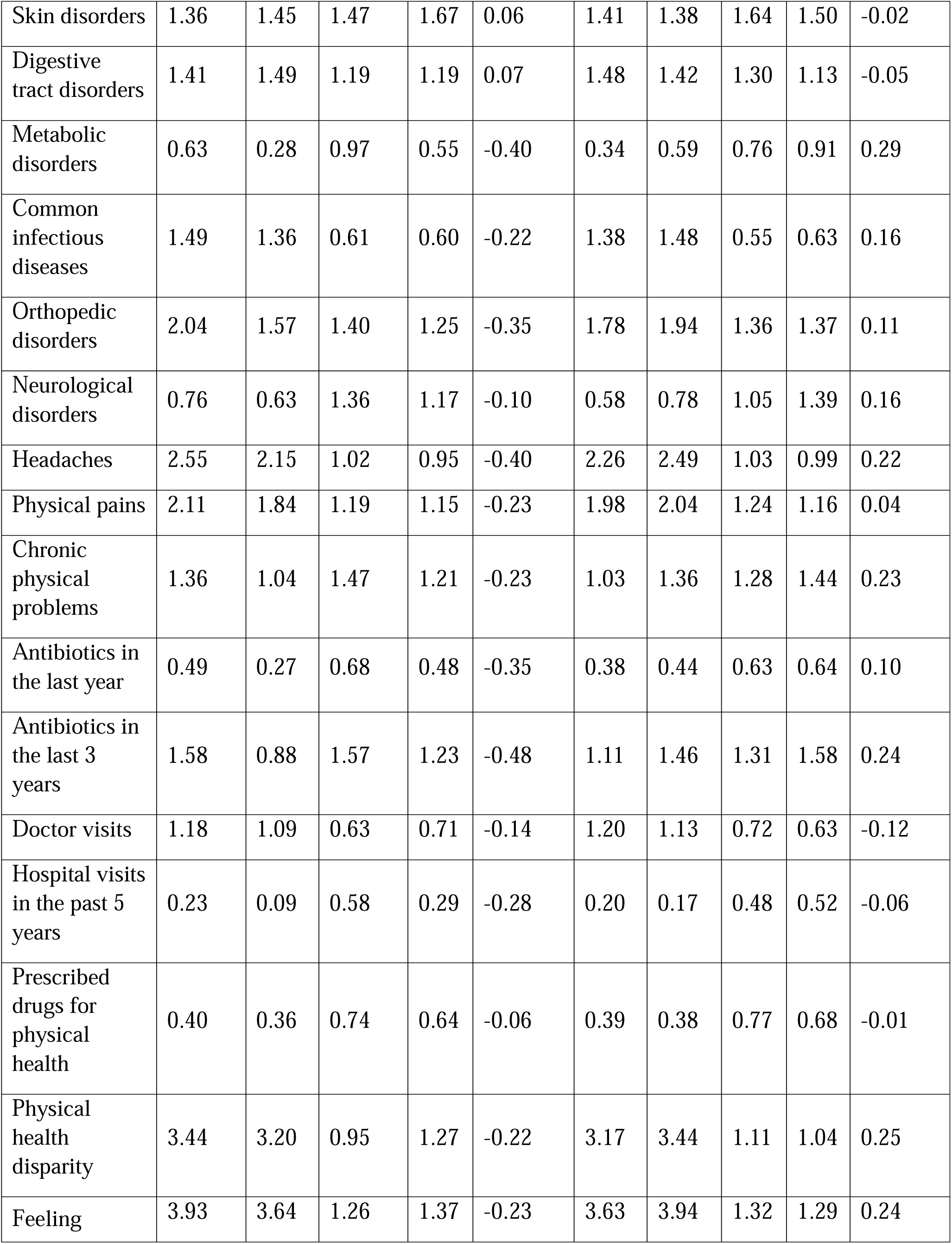
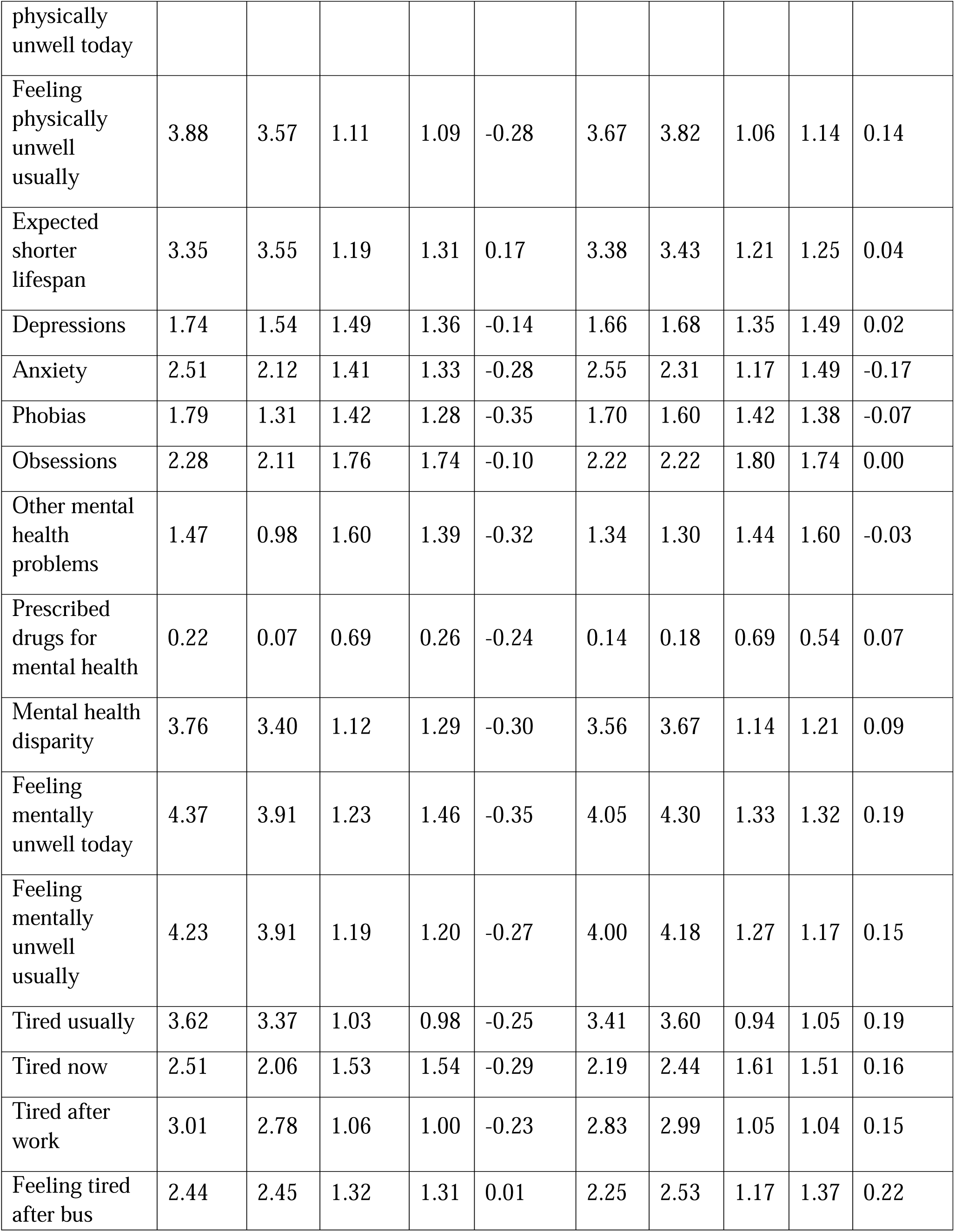
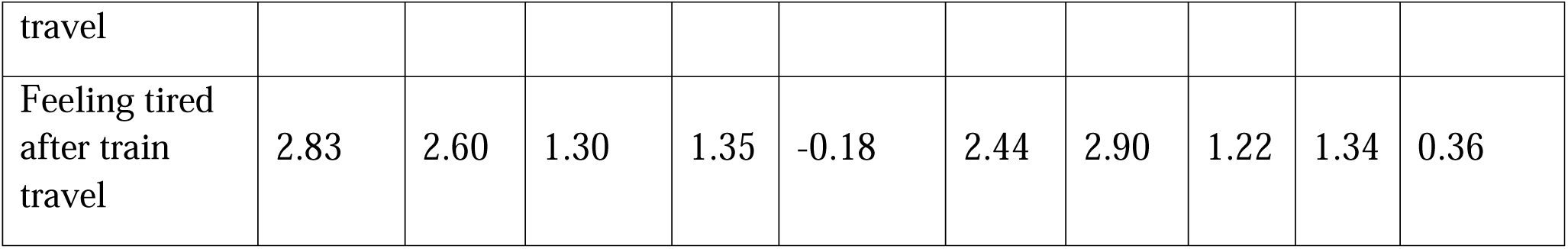
Descriptive statistics for the dependent variables.

**Supplementary Table S2.**
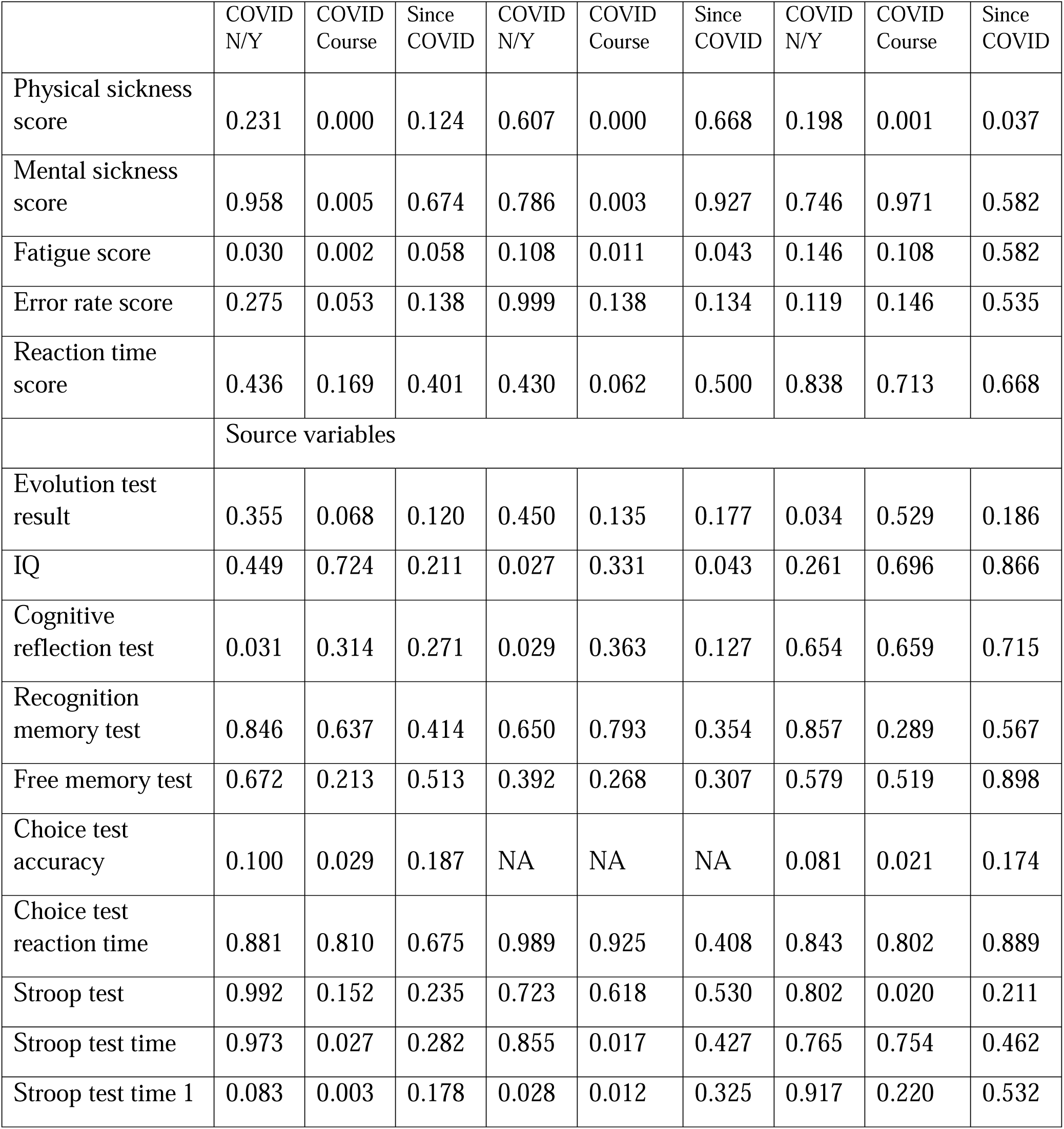
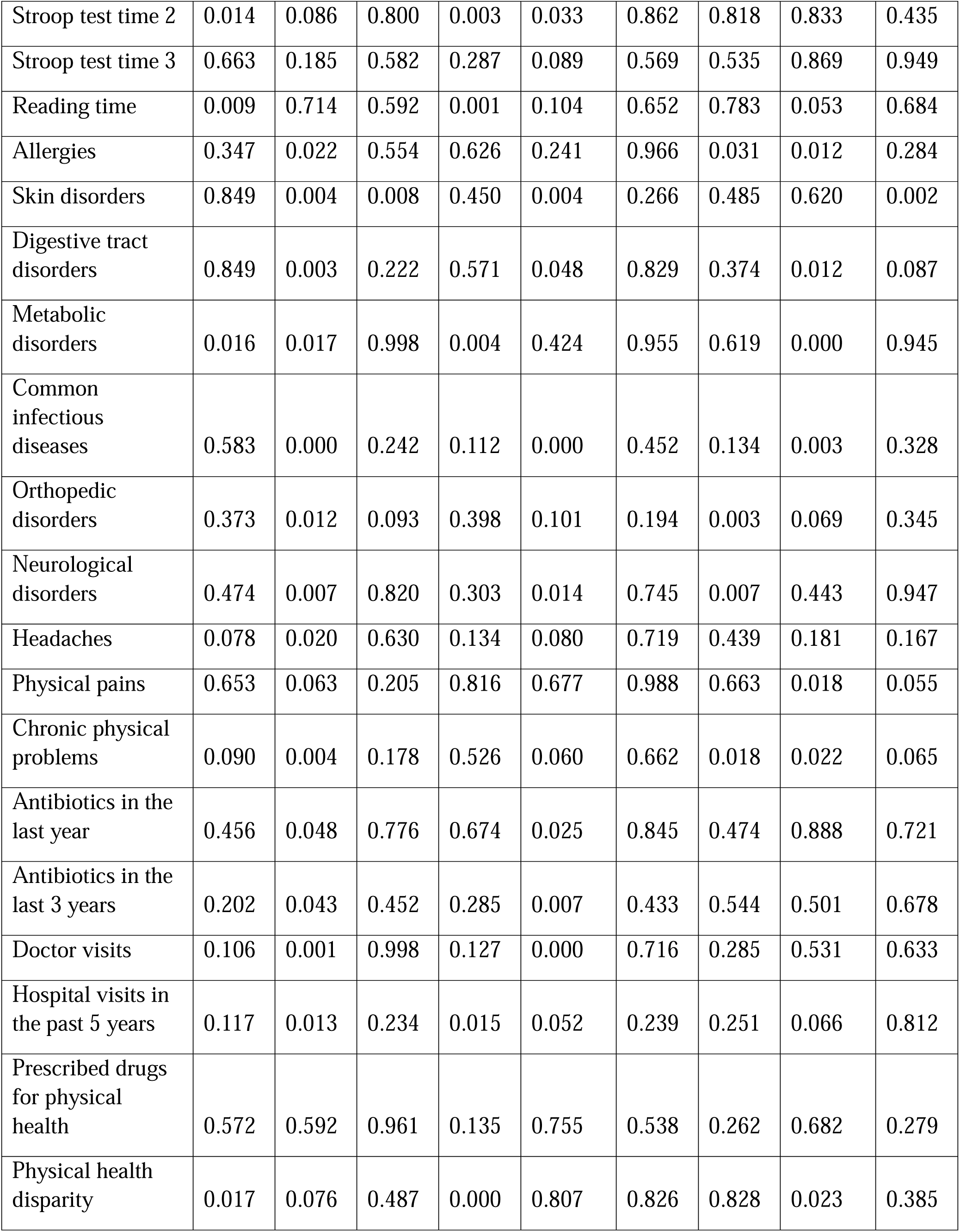
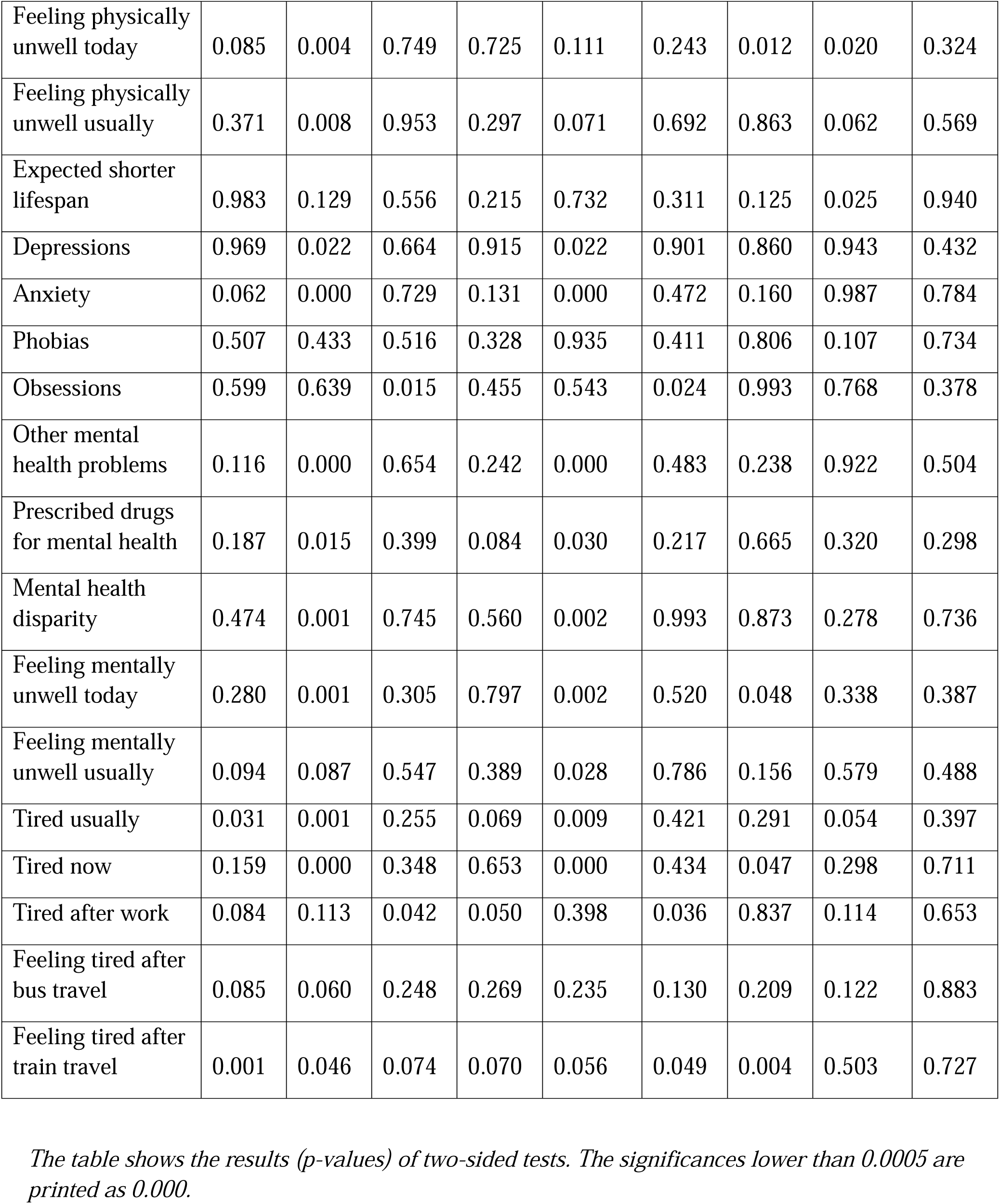
Significance of effects of COVID-19 exposure, COVID-19 course, and time since COVID-19 on health, performance, and fatigue.

**Supplementary Figure S1.**
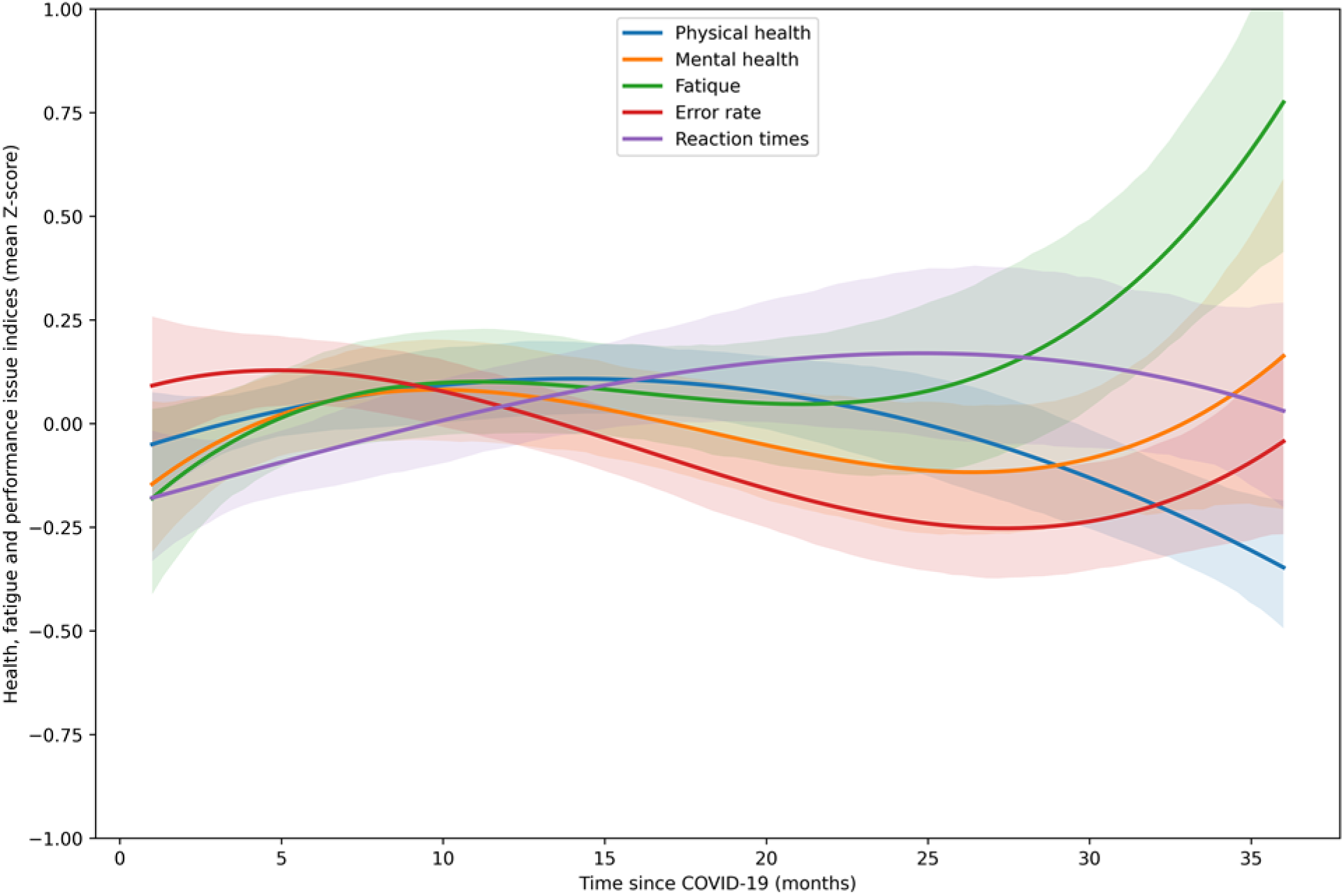
Third-degree polynomial trajectories of the disease course for health, performance, and fatigue over a span of three years. This scatterplot illustrates the relationships between five health-related variables (Physical Health, Mental Health, Fatigue, Error Rate, and Reaction Times) and the time elapsed since contracting COVID-19. Each variable is represented by a unique color, with data points fitted by a third-degree polynomial curve to visualize the trends. The bands around the lines represent 80% Confidence Intervals

